# Determining the optimal strategy for reopening schools, work and society in the UK: balancing earlier opening and the impact of test and trace strategies with the risk of occurrence of a secondary COVID-19 pandemic wave

**DOI:** 10.1101/2020.06.01.20100461

**Authors:** J. Panovska-Griffiths, C.C. Kerr, R.M. Stuart, D. Mistry, D.J. Klein, R.M. Viner, C. Bonell

## Abstract

**Background:** In order to slow down the spread of SARS-CoV-2, the virus causing the COVID-19 pandemic, the UK government has imposed strict physical distancing (‘lockdown’) measures including school ‘dismissals’ since 23 March 2020. As evidence is emerging that these measures may have slowed the spread of the pandemic, it is important to assess the impact of any changes in strategy, including scenarios for school reopening and broader relaxation of social distancing. This work uses an individual-based model to predict the impact of a suite of possible strategies to reopen schools in the UK, including that currently proposed by the UK government.

**Methods:** We use Covasim, a stochastic agent-based model for transmission of COVID-19, calibrated to the UK epidemic. The model describes individuals’ contact networks stratified as household, school, work and community layers, and uses demographic and epidemiological data from the UK. We simulate a range of different school reopening strategies with a society-wide relaxation of lockdown measures and in the presence of different non-pharmaceutical interventions, to estimate the number of new infections, cumulative cases and deaths, as well as the effective reproduction number with different strategies. To account for uncertainties within the stochastic simulation, we also simulated different levels of infectiousness of children and young adults under 20 years old compared to older ages.

**Findings:** We found that with increased levels of testing of people (between 25% and 72% of symptomatic people tested at some point during an active COVID-19 infection depending on scenarios) and effective contact-tracing and isolation for infected individuals, an epidemic rebound may be prevented across all reopening scenarios, with the effective reproduction number (R) remaining below one and the cumulative number of new infections and deaths significantly lower than they would be if testing did not increase. If UK schools reopen in phases from June 2020, prevention of a second wave would require testing 51% of symptomatic infections, tracing of 40% of their contacts, and isolation of symptomatic and diagnosed cases. However, without such measures, reopening of schools together with gradual relaxing of the lockdown measures are likely to induce a secondary pandemic wave, as are other scenarios for reopening. When infectiousness of <20 year olds was varied from 100% to 50% of that of older ages, our findings remained unchanged.

**Interpretation:** To prevent a secondary COVID-19 wave, relaxation of social distancing including reopening schools in the UK must be implemented alongside an active large-scale population-wide testing of symptomatic individuals and effective tracing of their contacts, followed by isolation of symptomatic and diagnosed individuals. Such combined measures have a greater likelihood of controlling the transmission of SARS-CoV-2 and preventing a large number of COVID-19 deaths than reopening schools and society with the current level of implementation of testing and isolation of infected individuals.

**Research in Context:** *Evidence before this study:* Since the onset of COVID-19 pandemic, mathematical modelling has been at the heart of informing decision-making, including the imposing of the lockdown in the UK. As countries are now starting to plan modification of these measures, it is important to assess the impact of different lockdown exit strategies including whether and how to reopen schools and relax other social distancing measures.

*Added value of this study:* Using mathematical modelling, we explored the impact of strategies to reopen schools and society in the UK, including that currently proposed by the UK government. We assessed the impact of opening all schools fully or in a phased way with only some school years going back, with a society-wide relaxation of lockdown measures and in the presence of a different levels of implementation of test-trace-isolate strategies. We projected the number of new COVID-19 infections, cumulative cases and deaths, as well as the temporal distribution in the effective reproduction number (R) across different strategies. Our study is the first to provide quantification of the amount of testing and tracing that would be needed to prevent a second wave of COVID-19 in the UK under different reopening scenarios. To account for uncertainties within the stochastic simulation, we also simulated different levels of infectiousness of children and young adults under 20 years old compared to older ages.

*Implications of all the available evidence:* Evidence to date points to the need for additional testing, contact tracing, and isolation of individuals who have either been diagnosed with COVID-19, or who are considered to be at high risk of carrying infection due to their contact history or symptoms. Our study supports these conclusions and provides additional quantification of the amount of testing and tracing that would be needed to prevent a second wave of COVID-19 in the UK under different lockdown exit strategies. Reopening schools and society alongside active testing of the symptomatic population (between 25% and 72% of people with symptomatic COVID-19 infection depending on scenarios) and with an effective contact-tracing and rapid isolation of symptomatic and diagnosed individuals, will not only prevent a secondary pandemic wave, but is also likely to be able to control the transmission of SARS-CoV-2, via keeping the R value below 1, thus preventing a large number of COVID-19 cases and deaths. However, in the absence of fully implemented large-scale testing, contact-tracing and isolation strategy, plans for reopening schools, including those currently proposed by the UK government, and the associated increase in work and community contacts, are likely to induce a secondary pandemic wave of COVID-19.

## Introduction

The COVID-19 pandemic, caused by the novel coronavirus SARS-CoV-2 virus, continues to spread globally with more than 6 million reported cases and over 360,000 deaths worldwide as of 31 May 2020. In the UK, since the first two reported cases on 31 January 2020 and the first reported COVID-19-related death on 7 March 2020, the number of reported cases and deaths has been increasing steadily with over 272,000 reported cases and over 38,000 deaths reported up to 31 May 2020.

To slow down the virus spread, flatten its epidemic curve, reduce the morbidity and mortality of the pandemic, and not overwhelm the National Health Service (NHS), the UK government imposed strict physical distancing (‘lockdown’) measures on 23 March 2020. Such mitigation strategies aimed to reduce the contact rates across the population, and consequently reduce the number of secondary infections quantified by the effective reproduction number in the population R. For epidemic control, R needs to be less than 1, so that the number of new infections is lower than the number of recovered infections allowing the epidemic to remain on the declining part of the epidemic curve.

Informed by evidence from previous influenza epidemics and by mathematical modelling of the potential spread and mortality of this pandemic^2^, and following the example of the countries affected earlier^3^, schools closures have occurred worldwide as a key element of COVID-19 lockdown measures. On 19 March 2020, UNESCO estimated that 1.6 billion children and young people across >180 countries had stopped attending school.^3^ In the UK, schools for 4-18 years old have been ‘dismissed’ rather than completely closed, remaining open for the children of key workers and children with defined health, education, or social needs, though uptake has been estimated to be only around 2% of school children attending during lockdown.

While closing schools does reduce the contact rate within the population and hence reduces onward transmission, considerable harms arise from school closures.^4^ These include hampering health care and other key workers’ ability to go to work^4^; great loss of economic productivity^5^; and considerable damage to children and young people’s education, development, and physical and mental health^6-8^ arising from social isolation^9^, reduced social support and increased exposure to violence at home.^10^ These harms will inevitably be greater in poorer families, exacerbating inequalities.

Since the beginning of May, the rate of increase in the number of COVID-19-related hospitalisations and deaths in the UK has been less than earlier in the epidemic.^10^ As a consequence, the first steps have been made towards planning how the UK will exit the lockdown. For planning, it is important to assess whether the number of infections will increase again when lockdown measures are lifted, leading to a secondary COVID-19 pandemic wave.

As one of several aspects of exiting the lockdown measures, the UK government is currently aiming to reopen schools in a phased manner with the precise timetable to be determined by trends in infections, availability of community testing and other criteria. Students in reception, year one and year six in English primary schools are currently planned to return from 1 June 2020, followed soon after by other years of primary school. After that, the intention is for secondary school students to return, starting with those in year 10 and year 12 in July followed by other secondary school students in the following September. These options are based on assumptions of lower transmission among primary school children, as noted above, and on findings from early population testing suggesting very low COVID-19 infection or asymptomatic carriage rates, particularly in those under 10 years.^12^ Reopened schools might aim to incorporate physical distancing of 2 metres between students in the same classes,^13^ and disrupt contacts between students in different classes and years by staggering school start, break, lunch and end times. Introduction of proven infection control measures within schools, including cleaning and hand hygiene are also very likely to contribute. Reactive school closures are also planned in response to local outbreaks. In Taiwan, individual schools are closed when infections among two or more teachers or students are confirmed, and all schools in a city are closed when one third of individual schools are closed due to infection.^11^

Decisions about reopening schools while ensuring physical distancing and other measures to minimise COVID-19 transmission need to be informed by their impact, and to allow schools sufficient time and resources to plan and implement changes to timetabling, physical environments and support for teachers and vulnerable students. It is therefore critically important to understand the extent to which different options for reopening schools will maintain control of COVID-19 transmission or lead to secondary pandemic waves. This is particularly challenging because of the uncertainty about the importance of children and young people in COVID-19 transmission and the impact of school closures in COVID-19 control.^7^ While existing modelling has suggested that school closures contribute to prevention alongside other physical distancing interventions^2^; this generally assumes that transmissibility among children and young people is equivalent to that among adults. Data on susceptibility to and transmission of COVID-19 among children and adolescents are sparse.^5^ A population-based contact-tracing data on transmission in schools in Australia identified two likely secondary cases from 18 index cases and 863 contacts.^6^ Yet others have suggested that the attack rate is similar to that in adults^7^, and much of the data on school transmission comes from periods when schools have been fully or partially closed. A recent meta-analysis by some of us suggested that susceptibility to SARS-CoV-2 amongst children and adolescents was around half of that amongst adults.^8^ Early data on viral load suggest that symptomatic children may have similar COVID-19 viral load to adults^9^; however symptoms are much less common in children than adults and the degree of asymptomatic transmission by children is unknown.

In this paper, we use modelling to explore the impact of a suite of strategies to reopen schools combined with society-wide relaxing of the social distancing measures in the UK. Specifically, we aim to explore the impact of strategies to reopen schools fully in June or in September, in comparison to reopening schools in a phased gradual way as outlined by the UK Prime Minister on 10 May 2020, combined with and without increase in contact rate within community and workplaces. We conduct sensitivity analyses to assess how our results would change if under 20 years old are less infectious than older ages. The strategies we have explored have been discussed with members of scientific advisory bodies in the UK.

## Methods

### Transmission model

We modelled the spread of COVID-19 using Covasim, a stochastic agent-based model of SARS-CoV-2 transmission across a population. The model was developed by the Institute for Disease Modelling, and development and implementation details can be found at http://docs.covasim.org. Further details of the mathematical approach used for Covasim are contained in Kerr et al.^12^ Briefly, within the model, individuals were modelled as either susceptible to the virus, exposed to it, infected, recovered or dead. In addition, infected and infectious individuals were categorised as either asymptomatic or in different symptomatic groups: pre-symptomatic (before viral shedding has begun) and with mild, severe or critical symptoms. A schematic of the model is given in Figure 1. The code used to run all simulations contained in this paper is available from https://github.com/Jasminapg/Covid-19-Analysis, while the model code is available from https://github.com/InstituteforDiseaseModeling/covasim.

**Figure 1.**
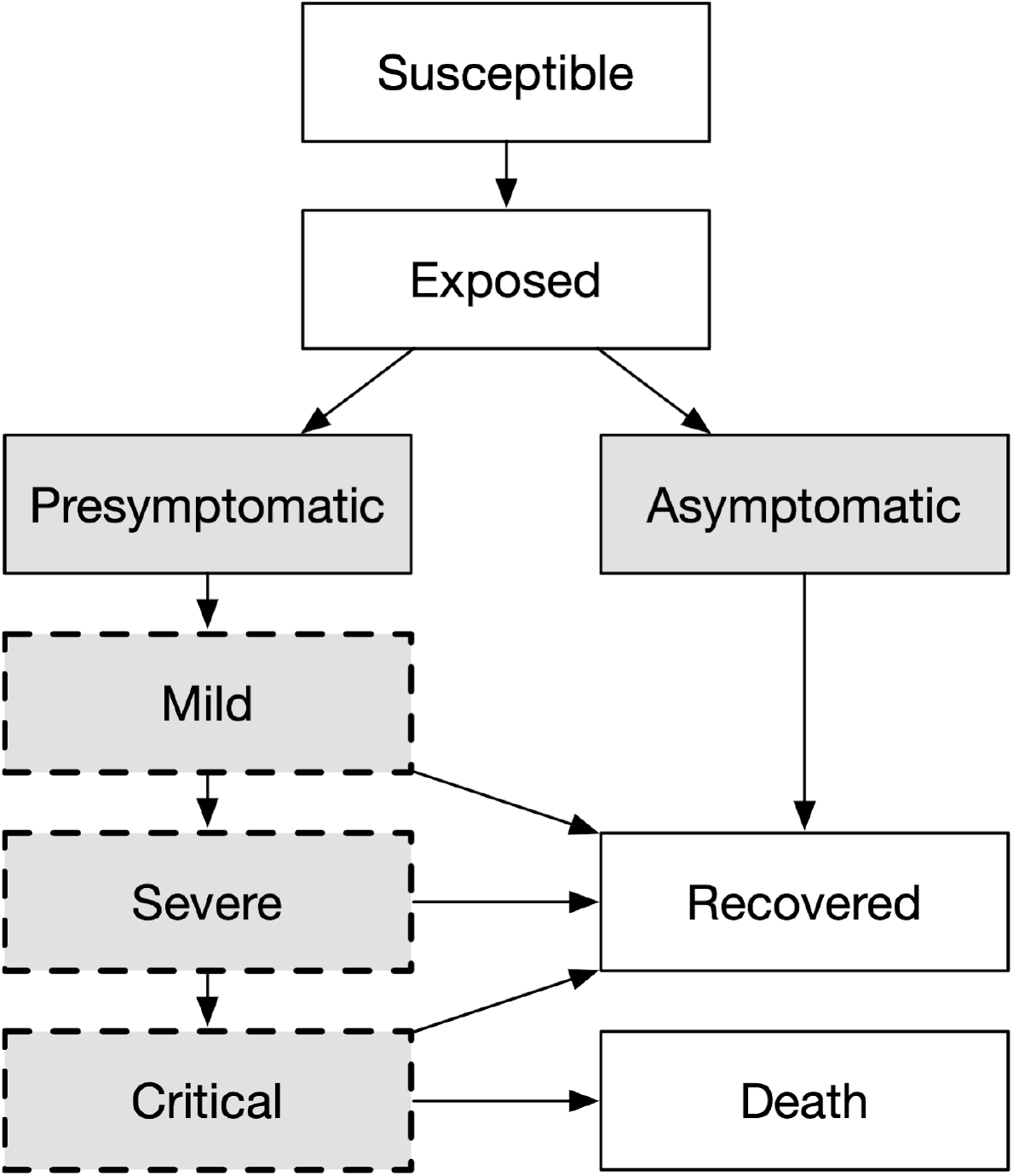
Modelled disease states. Grey shading indicates that an individual is infectious and can transmit the disease to other susceptible individuals. States with a dashed border are considered to be symptomatic for the purpose of testing eligibility with TI and TTI strategies. This schematic is reproduced from existing work from this group in Kerr et al.^12^

Covasim’s default parameters determine the ways in which people progress through the states depicted in Figure 1, including the probabilities associated with onward transmission and disease progression, duration of disease by acuity, and the effects of interventions; these were collated during Covasim’s development and summarise the evidence available up until May 10, 2020.^12^ In addition, Covasim is pre-populated with demographic data on population age structures and household sizes by country, and uses these to generate population contact networks for the setting. To apply Covasim to model the epidemic in the UK, we used Covasim’s inbuilt defaults to generate a population of 100,000 agents with contact networks across schools, workplaces, household and community. We then seeded 4,500 cases in the population on 21/01/2020 and adjusted the per-contact transmission probabilities during the calibration process. The number of seeded infectious individuals was varied during calibration and the final number was chosen to reflect the epidemic trend to date and consistent with undetected community transmission as well as possible multiple importation events.

Within the model, susceptible individuals come into contact with infectious individuals, and transmission of the virus occurs with a daily probability *β*. We assumed that *β* varies with the nature of contacts; we defined household, school, workplace and community contacts as different contact groups. With this assumption, imposing lockdowns would reduce the school, work and community mixing and hence reduce the equivalent *β* for these groups. This approach is similar to that in Ferguson et al,^2^ one of the studies that directly influenced the imposition of lockdown measures in the UK. Currently in the literature, there is considerable uncertainty about whether *β* is age-dependent^13^ or differs across asymptomatic and symptomatic cases. Therefore, during the calibration process, we varied *β* to match the UK epidemic trend to date, and specifically, reported cumulative deaths from COVID-19 and reported number of COVID-19 infections between 21/01/2020 and 20/05/2020 (see supplementary material).

We simulated a range of different school reopening scenarios (see below) in combination with society-wide relaxing of some lockdown measures and estimated the number of new infections and cumulative cases over time until 31 May 2021. Across different strategies, we calculated the number of cumulative infections and recoveries, number of new infections and cumulative deaths as well as the timeseries of the effective reproduction number R.

### School and society reopening scenarios

The UK Prime Minister announced on 10^th^ May 2020 that as one of several aspects of exiting the lockdown measures, the UK government is currently aiming to reopen schools in a phased manner. The current target is that, from 1 June 2020, students in reception (children first entering school), year one and year six in English primary schools would return, followed in July by other years of primary school. After that, secondary school students would return, starting with those in year 10 and year 12 in July followed by other secondary school students the following September. This timetable is dependent on various criteria, including reduced infections and the implementation of community test, trace and isolate (TTI) strategies, so delays are possible. Therefore, a second plausible scenario is that students in reception, year one and year six would return on 1 July 2020, followed by all other primary and secondary school students on 1 September 2020. We explore two further scenarios in order to provide a benchmark against which to compare the first two scenarios. Our third scenario is that all schools would reopen to all students on 1 June 2020 and our fourth that all schools would reopen to all students on 1 September 2020. Details of the scenarios are contained in Table 1.

**Table 1:**
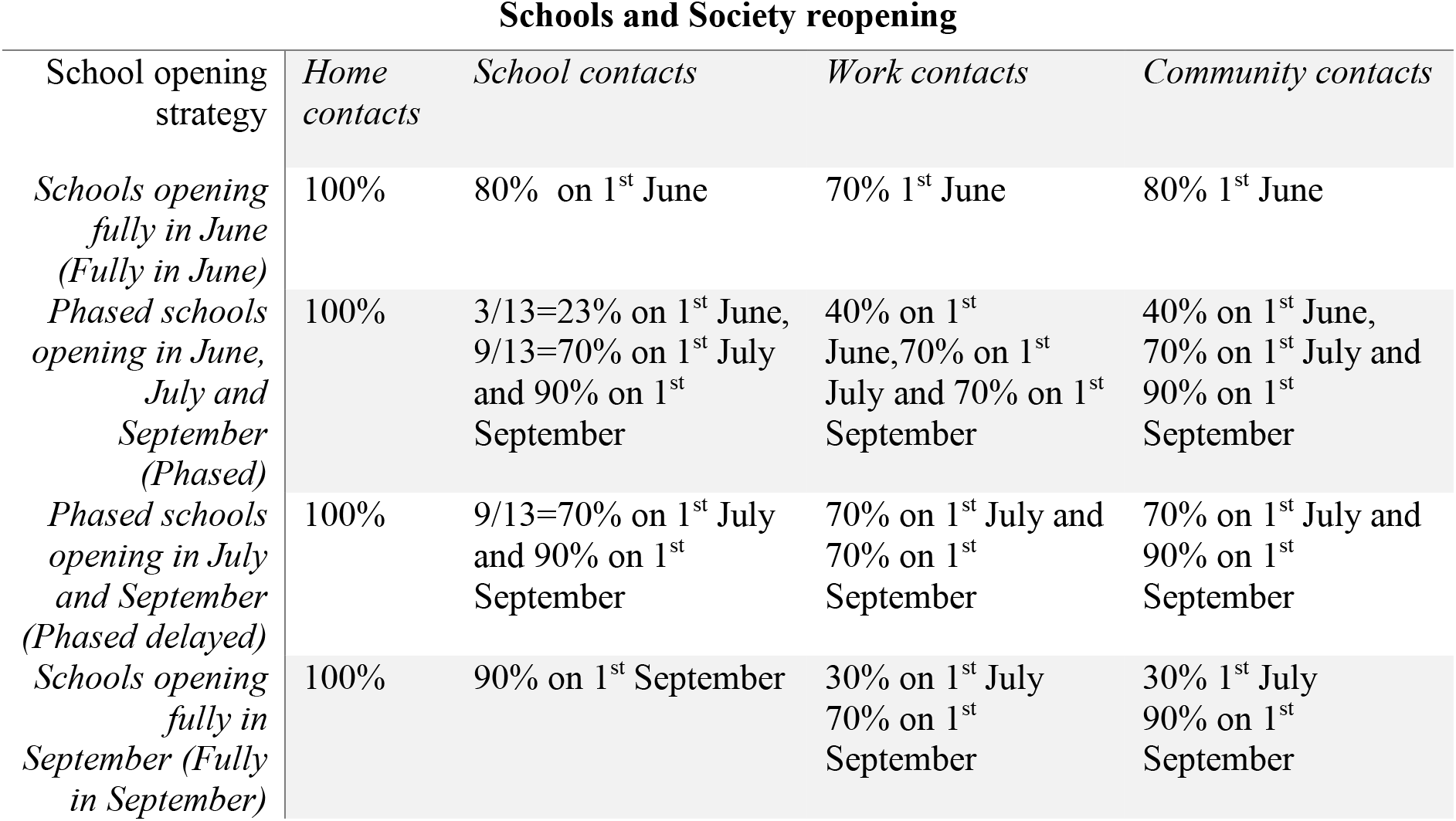
Description of strategies to reopen schools, workplace and society simulated in the model. Each intervention is simulated by altering the daily transmission probability due to home, school, work and/or community contact with details presented in the supplementary material.

| Schools and Society reopening |  |  |  |  |
| --- | --- | --- | --- | --- |
| School opening strategy | Home contacts | School contacts | Work contacts | Community contacts |
| *Schools opening fully in June (Fully in June)* | 100% | 80% on 1<sup>st</sup> June | 70% 1<sup>st</sup> June | 80% 1<sup>st</sup> June |
| *Phased schools opening in June, July and September (Phased)* | 100% | 3/13=23% on 1<sup>st</sup> June, 9/13=70% on 1<sup>st</sup> July and 90% on 1<sup>st</sup> September | 40% on 1<sup>st</sup> June, 70% on 1<sup>st</sup> July and 70% on 1<sup>st</sup> September | 40% on 1<sup>st</sup> June, 70% on 1<sup>st</sup> July and 90% on 1<sup>st</sup> September |
| *Phased schools opening in July and September (Phased delayed)* | 100% | 9/13=70% on 1<sup>st</sup> July and 90% on 1<sup>st</sup> September | 70% on 1<sup>st</sup> July and 70% on 1<sup>st</sup> September | 70% on 1<sup>st</sup> July and 90% on 1<sup>st</sup> September |
| *Schools opening fully in September (Fully in September)* | 100% | 90% on 1<sup>st</sup> September | 30% on 1<sup>st</sup> July 70% on 1<sup>st</sup> September | 30% 1<sup>st</sup> July 90% on 1<sup>st</sup> September |

To implement these scenarios within Covasim, we adjusted the transmission probability for household, school, workplace and community contacts, increasing it proportionally to the number of school years going back. For example, consider the phased reopening of schools scenarios where reception, year one and year six only go back to school on 1 June 2020, followed by years two to five plus years 10 and 12 going back on 1 July 2020 and then all school years going back on 1 September 2020. This was implemented in the model by increasing the school transmission probability from 2% during the lockdown to 3/13=23% on 1 June 2020, to 9/13=70% on 1 July 2020 and then to 90% on 1 September 2020. We assumed 90% rather than 100% transmission probability in September to account for protective measures (e.g. wearing masks, keeping a 2m distance at all time etc) assumed to be put in place if a large cohort of students goes back to school at one time. The exact % changes to the transmission probability across scenarios are listed in Table 1.

Across scenarios, we assumed that with increase in school transmission probability, workplace and community transmission probabilities would also increase respectively, to account for a) increased social mixing with reopening of schools and b) relaxation of social distancing restrictions on work, leisure and community activities that are occurring alongside school reopening. Details of the exact model assumptions regarding these scenarios can be found in the supplementary material. In this paper we focus on the differences in school reopening strategies and the degree of increase in broader societal social mixing these would allow due to parents returning to work.

### Testing, tracing and isolation strategies

In line with current policy in the UK, we also modelled the implementation of strategies to test those in the population presenting with COVID-19-like symptoms and isolate those testing positive; this is a test-isolate or TI strategy. This has been the strategy in the UK since 23 March 2020. Starting on 1 June 2020, in line with plans underway in the UK, we also simulated a strategy to trace contacts of those people who test positive to infection. This test-trace-isolate (TTI) strategy aims to encourage testing of symptomatic individuals and trace and isolate their contacts that are symptomatic or diagnosed positive. The TTI strategy is simulated in the model by increasing the level of symptomatic testing from 1 June 2020 and introducing two coverage levels of tracing. Firstly, to resemble a pessimistic scenario for tracing capability, we modelled a tracing coverage of 40%, and secondly, to resemble an optimistic scenario, we modelled a tracing coverage of 80%.

We examined four scenarios of schools opening, each modelled with three testing strategies: the TI strategy and the TTI strategy assuming either 40% or 80% tracing of contacts of positive diagnosis. Details of the specific model changes to implement TI and TTI are contained in the supplementary material. Briefly, in the model we accounted for testing strategies by specifying the probabilities with which people with different symptoms receive a test each day. For both testing strategies, we assumed that people who present themselves with symptoms will be tested with a daily probability that was determined in the calibration and we also included a small testing of asymptomatic people based on testing of NHS staff and other key essential workers such as careers at care homes. Assuming an average symptomatic period of roughly 10 days, these can be translated to a measure of the testing rate for people with COVID-19 infections being tested at some point during their illness. During our calibration (supplementary material) we predicted that under the current strategy the daily probability of testing a symptomatic person is 1.2% and an asymptomatic person is 0.057%, corresponding to about 11% of people with symptomatic and 0.7% with asymptomatic COVID-19 infections being tested at some point during their illness. With the TTI strategy, from 1 June 2020, we assumed that this testing of symptomatic and asymptomatic people continued, but was supplemented by a strategy to trace their contacts with coverage of 40% or 80%. We used the model to derive a testing level necessary to avoid the secondary pandemic wave with these two tracing strategies. With both strategies, TI and TTI, we assumed a delay of one day to receive the test result and once an individual tested positive, they were immediately isolated for 14 days. In the model, this isolation reduced their infectiousness by 90%. In addition, with both strategies, symptomatic people were also isolated with their infectiousness reduced by 50%. More details are available in the supplementary material.

### Varying the infectiousness of under 20 years old compared to other ages

Given uncertainties about the role of different age groups in transmission,^5^ we explored how varying the transmission among children and young people compared to adults would alter our predictions. Within the simulation, we achieved this by changing the infectiousness of anyone under 20 years old to be 50%,^14^ or 100% of the infectiousness of adults. Our primary analysis assumed transmission to be 100% of that of adults, with 50% being a sensitivity analysis. In both cases we calibrated the model to the UK epidemic, matching the number of reported cases and deaths until 20^th^ May 2020.

## Results

The outcomes from our simulations are shown in Figures 2-4. Figure 2 contains the projections of the number of new COVID-19 infections, while Figure 3 shows the deaths associated with COVID-19 over time since the onset of the pandemic until 31 May 2021 across the different scenarios considered. Figure 4 shows the temporal profiles of the effective reproduction number R, across all twelve scenarios. Finally, Figure 5 presents the cumulative number of COVID-19 infections and associated deaths across all scenarios.

**Figure 2:**
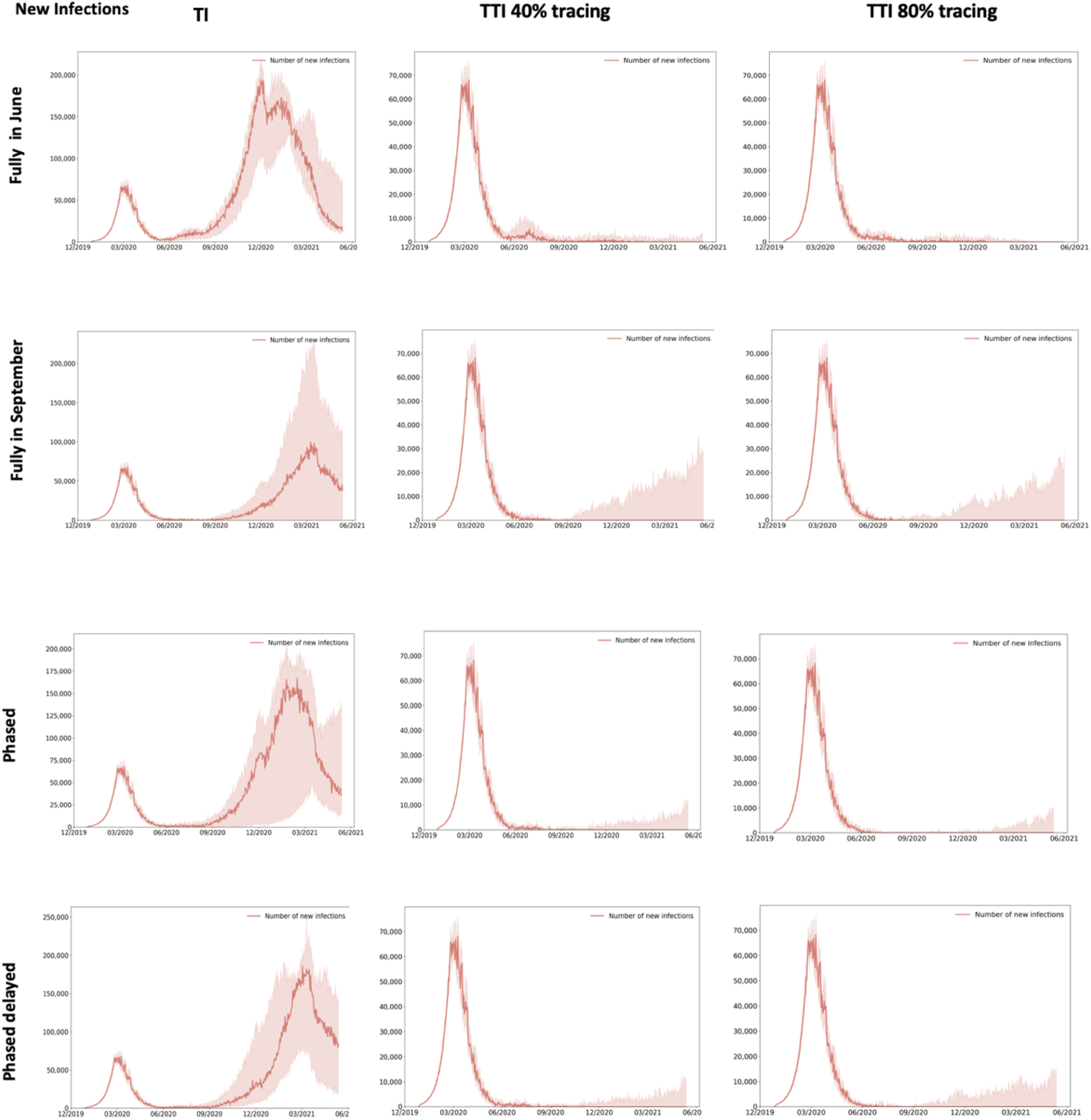
Model forecasting of new COVID-19 infections across different school and society reopening scenarios in presence of different test-isolate (TI) and test-trace-isolate (TTI) strategies. Each TTI strategy is simulated assuming a level of either 40% (TTI 40% tracing) or 80% (TTI 80% tracing) tracing of contacts of positive diagnoses. Across tiles in this figure we present the temporal distribution of new COVID-19 infections between 21/02/2020 and 31/05/2021 showing the median across six^2^ simulations as the solid red line and the simulation noise (as a measure for uncertainty across different simulations) as the red-shaded part. Simulations were done using the Covasim model adapted with details given in the supplementary material, and parameters simulated across scenarios listed in Tables 1-2.

**Table 2:** Description of test-isolate (TI) and test-trace-isolate (TTI) strategies simulated in the model. Each TTI strategy is simulated assuming a level of either 40% (TTI 40% tracing) or 80% (TTI 80% tracing) tracing of contacts of positive diagnoses. Testing with TI and TTI strategies is simulated by specifying the probabilities with which people with different symptoms receive a test each day. In the model this was implemented by setting daily probability of testing an asymptomatic person (*p_as_ =* 0.00075) and of a symptomatic person (*p*_s_) with TI, TTI 40% tracing and TTI 80% tracing. Before June 2020, *p_s_* = 0012 was determined during the calibration and used for TI forecasting, while after June 2020 we determined a minimum *p_s_* to avoid a secondary COVID-19 wave across different scenarios (2^nd^ column). The latter values were used for TTI 40% tracing and TTI 80% tracing forecasting. The proportion of the population with symptomatic COVID-19 infection (3^rd^ column) is derived from the equation 1 − (1 − *p_s_*)*^T^* where *T* is the average infectiousness period assumed to be roughly 10 days. Tracing of contacts is modelled with some delay (4^th^-7^th^ column) and under the assumption that the infected individuals are immediately quarantined for 14 days (8^th^ column).

| Test-Trace-Isolate strategies | Daily probability of testing<sup>1</sup> in the model across scenarios($p_s$) | Proportion of people with symptomatic COVID-19 infections being tested at some point during their illness | Delay in contact tracing (days) | | | | Isolation duration |
| --- | --- | --- | --- | --- | --- | --- | --- |
|  |  |  | Home contacts | School contacts | Work contacts | Community contacts |  |
| TI | 0.012 | 11% | 0 | 1 | 1 | 2 | 14 days |
| TTI 40% tracing | 0.052-0.12 | 41%-72% | 0 | 1 | 1 | 2 | 14 days |
| TTI 80% tracing | 0.029-0.09 | 25%-61% | 0 | 1 | 1 | 2 | 14 days |

**Figure 3:**
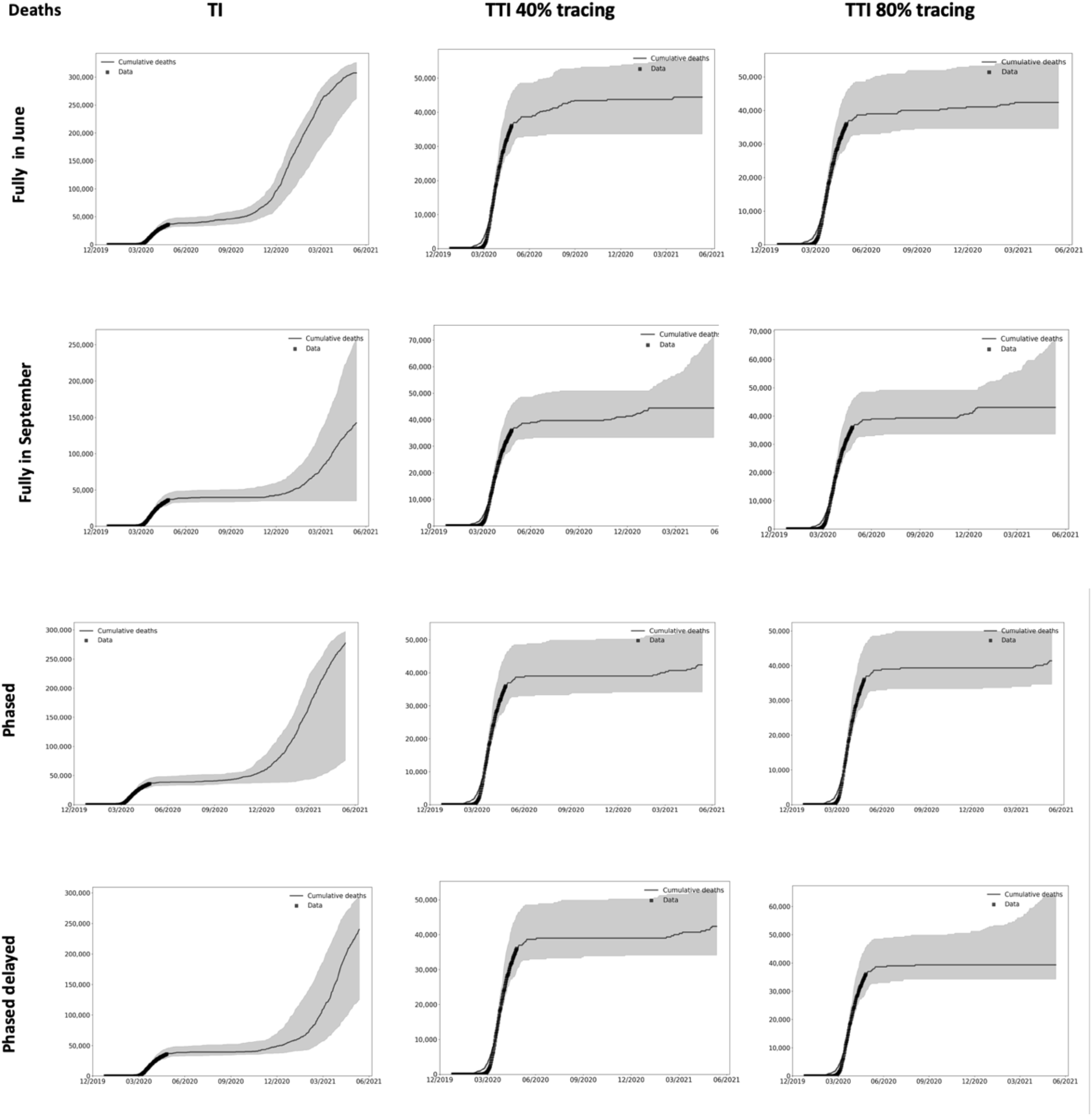
Model forecasting of cumulative COVID-19 deaths across different school and society reopening scenarios in presence of different test-isolate (TI) and test-trace-isolate (TTI) strategies. Each TTI strategy is simulated assuming a level of either 40% (TTI 40% tracing) or 80% (TTI 80% tracing) tracing of contacts of positive diagnoses. Across tiles in this figure we present the temporal distribution of deaths associated with COVID-19 between 21/02/2020 and 31/05/2021, showing the median across six simulations as the solid black line and the simulation noise (as a measure for uncertainty across different simulations) as the grey-shaded part. Simulations were done using the Covasim model adapted with details given in the supplementary material, and parameters simulated across scenarios listed in Tables 1-2.

**Figure 4:**
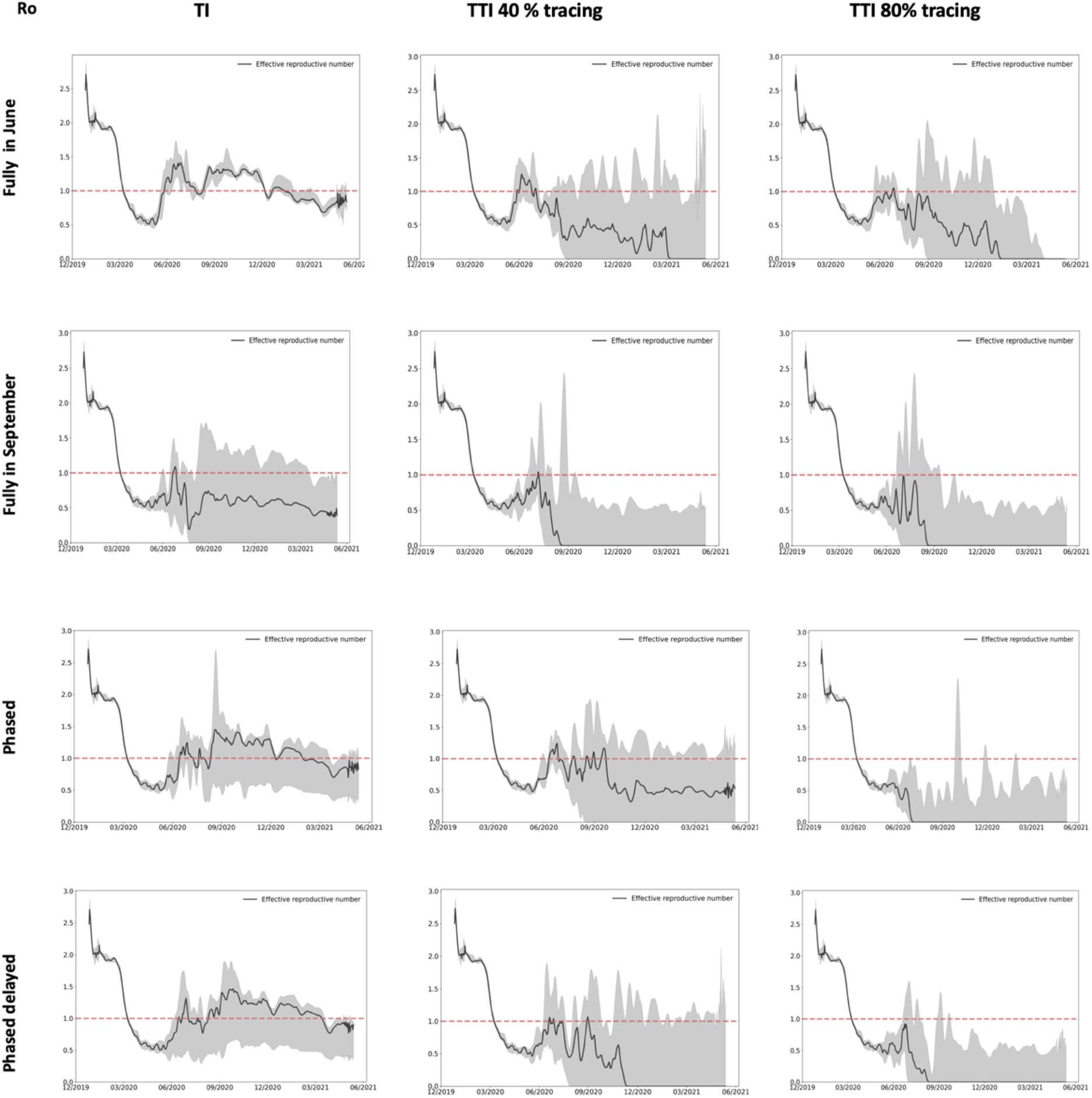
Model forecasting of effective reproduction number R over time across different school and society reopening scenarios in presence of different test-isolate (TI) and test-trace-isolate (TTI) strategies. Each TTI strategy is simulated assuming a level of either 40% (TTI 40% tracing) or 80% (TTI 80% tracing) tracing of contacts of positive diagnoses. Across tiles in this figure we present the temporal distribution of effective reproduction number R between 21/02/2020 and 31/05/2021, showing the median across six simulations as the solid black line and the simulation noise (as a measure for uncertainty across different simulations) as the grey-shaded part. Simulations were done using the Covasim model adapted with details given in the supplementary material, and parameters simulated across scenarios listed in Tables 1-2.

**Figure 5:**
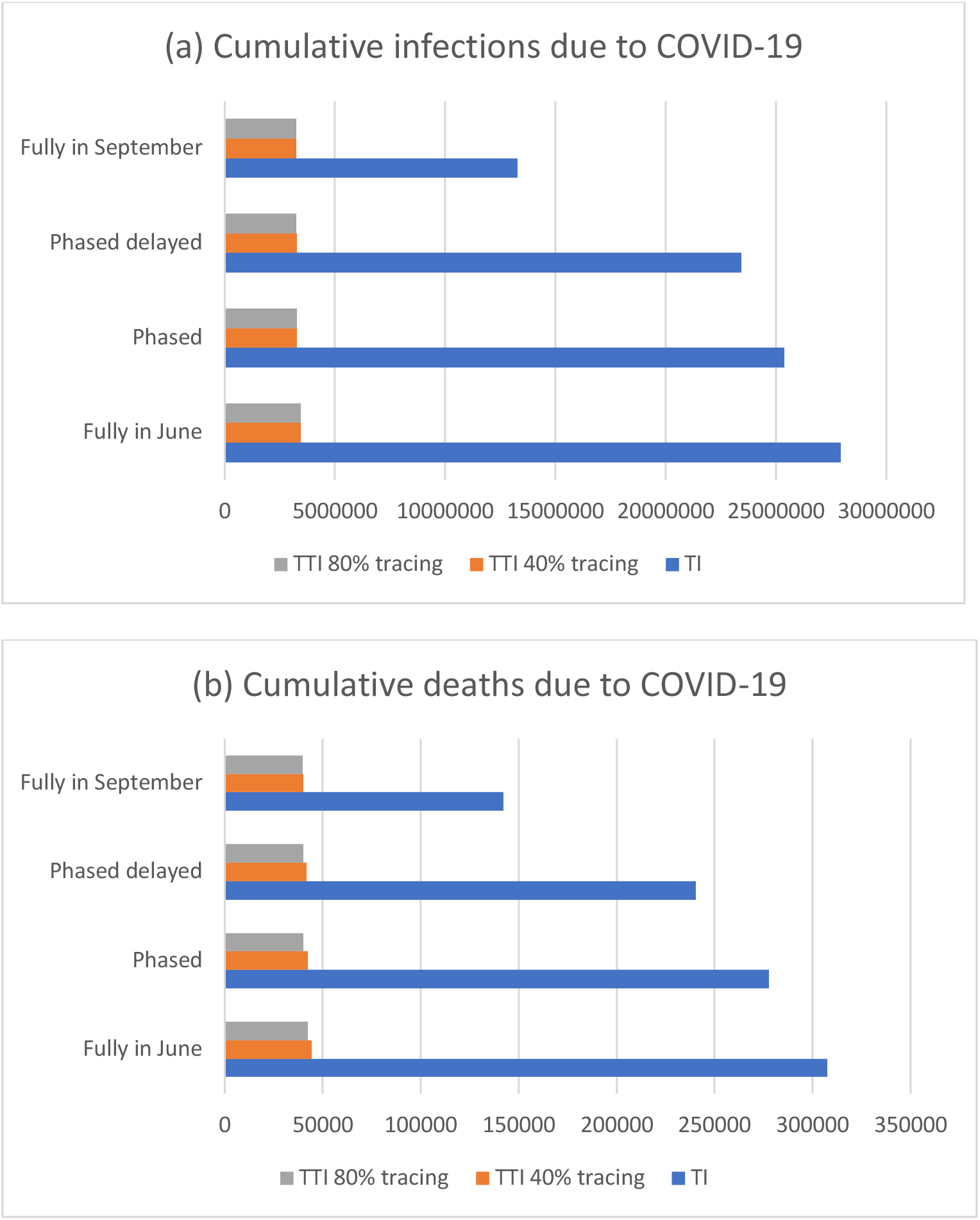
Model forecasting of overall cumulative COVID-19 infections (a) and deaths associated with COVID-19 (b) across different school and society reopening scenarios in presence of different test-isolate (TI) and test-trace-isolate (TTI) strategies. Each TTI strategy is simulated assuming a level of either 40% (TTI 40% tracing) or 80% (TTI 80% tracing) tracing of contacts of positive diagnoses. The numbers within these bar-charts are collated from respective tiles in Figure 2 (for part (a)) and Figure 3 (for part (b)).

### Impact of TTI strategies

Our findings suggest that it may be possible to avoid a secondary pandemic wave across all school reopening scenarios with an enhanced strategy that tests between 25% and 72% of people with symptomatic COVID-19 infection, traces 40%-80% of their contacts and isolates symptomatic cases and those with positive diagnosis (Figure 2-4, 2^nd^ column and 3^rd^ column). In this case, across scenarios, the predicted number of cumulative infections and deaths would be dramatically reduced (Figure 5). Across different scenarios of school and society reopening and different tracing levels, such a TTI strategy would need to test a sufficiently large proportion of the population with COVID-19 symptomatic infection to prevent a secondary wave. If we assume that contact tracing can achieve a coverage of 40% of the contacts of those testing positive for COVID-19 being traced and isolated if symptomatic or diagnosed positive, then a second wave could be prevented by testing the following proportions of those with symptomatic infection for each scenario: 72% where all schools return in June; 51% where schools return in phases from June; 46% where the phased return is delayed till July; and 43% where the phased return is from September. If we assume that 80% of contacts are traced, then the corresponding figures are less: 61%, 43%, 28% and 25%. With enhanced testing and effective tracing of their contacts, followed by isolation of symptomatic and diagnosed positive individuals, R will continue to decline until it eventually diminishes and infections are cleared (Figures 2-3, 3^rd^ column).

### Impact of TI strategies

If reopening of schools and society in the UK on 1 June were not to be accompanied by a TTI strategy, our results suggest that across all scenarios, a secondary pandemic wave would be likely (Figure 2-4 first column). The size of the predicted secondary wave of infections would be related to the proportion of school years returning to school and the associated proportion of societal reopening. Reopening both primary and secondary schools on 1 June 2020 or having phased reopening of schools on 1 June 2020 or 1 July 2020 and the rest of the year groups on 1 September 2020 would result in a slightly larger secondary wave of new infections and deaths than if schools reopen fully in September (Figures 2-3;1^st^ column). With schools opening fully or in phases in June or July this second wave would be around 2.2-2.5 times larger than the first COVID-19 wave in the UK. Reopening all schools in September would also produce a secondary wave but around 1.3 times larger than the first.

Across scenarios, the secondary COVID-19 wave would occur at different times. Specifically, a possible secondary wave of COVID-19 is predicted to occur earlier if schools reopen sooner at any capacity (Figure 2, 1^st^ column). Across the scenarios considered, the secondary wave is predicted to occur later if schools reopen later. For example if schools open in September, then a second COVID-19 wave may occur around March 2021, in comparison to if they open in June or July when the second pandemic wave may occur in December 2020 (Figure 2, 1^st^ column).

### R projections

The temporal profiles of the effective reproduction number R follow the trend of the time series of new infections (comparing respective tiles across Figure 2 and 4). R evidently increases over the threshold of 1, suggesting an increase in the number of new infections, when a secondary COVID-19 wave occurs (1^st^ column in Figures 2 and 4). However, R can also oscillate around 1, suggesting a small number of infected people and some infection still present, although not enough to cause a secondary wave. In this case, the implemented strategy could prevent a secondary wave but not fully eliminate infection (Figure 2-4, 2^nd^ column). Across all scenarios of school and society reopening and different tracing levels, the TTI strategy would need to test a sufficiently large proportion of the population with COVID-19 symptomatic infection and trace their contacts with sufficiently large coverage, for R to diminish and infection to be fully cleared (Figures 2-3 3^rd^ column). Specifically, our simulations predict that the time when R diminishes depends on the level of TTI; when tracing is lower and testing higher R diminishes quicker with schools reopening in September (2^nd^ column in Figure 4), while if testing is lower and tracing higher, R diminishes quickest with schools reopening in a phased manner from June or July or in September (3^rd^ column in Figure 4). The exact relationship between timing of R diminishment at different levels of TTI from June 2020 will be explored in subsequent analyses.

### Varying the infectiousness of under 20 years old compared to other ages

When we reran the model with infectiousness amongst under 20 years old assumed to be 50% of that among older ages, our results remained largely unchanged. To match the UK epidemic, with assumed reduced infectiousness among under 20 years old, we needed to increase the transmission probability and the testing level to date in the calibration (details in the supplementary material). The model, run under the same scenarios, suggests that it is possible to avoid a secondary COVID-19 wave across all scenarios of school and society reopening and different tracing levels, if the TTI strategy tests a sufficiently large proportion of the population with COVID-19 symptomatic infection and traces their contacts with sufficiently large coverage (Figure S3 of the supplementary material). This, alongside the impact of variable susceptibility of children will be explored further in subsequent analyses.

## Discussion

Our modelling results suggest that if schools and society reopened with a large-scale TTI strategy that tests between 25% and 72% of people with symptomatic COVID-19 infections and traces 40-80% of their contacts, with both dependent on the reopening scenario, a secondary COVID-19 wave may be prevented in the UK. With the current strategy of schools reopening in phases from 1 June 2020, prevention of a second wave would require testing of 51% of symptomatic COVID-19 infections and tracing of 40% of their contacts or testing of 43% of symptomatic COVID-19 infections and tracing of 80% of their contacts, combined with isolation of all symptomatic or diagnosed positive for infection individuals (Figures 2-3). In addition, such measures would markedly reduce cumulative numbers of new infections and deaths, and keep R below 1 (Figure 4). This is the case both in the main analyses assuming infectivity of under 20 years old is 100% of adults and when we assume that infectivity of under 20 years old is 50% that of adults (Figure S3 in supplementary material). We note that depending on the overall population prevalence of COVID-19-like illness, achieving this level of coverage with a TTI strategy would likely require testing a large number of people.

However, we also predict that in the absence of a sufficiently strong TTI strategy, reopening schools combined with accompanied reopening of the society across all scenarios, will induce a secondary COVID-19 wave. For example, our modelling results suggest that reopening both primary and secondary schools on 1 June 2020 without effective TTI would result in a rise in R above 1 and a resulting secondary wave of infections 2.5 times the size of the original COVID-19 wave.

Evidence from countries like South Korea^20,21^ where large-scale testing and contact-tracing have been able to control the spread of COVID-19, points to the need for additional testing, effective contact tracing, and isolation of individuals who have either been diagnosed with COVID-19, or who are considered to be at high risk of carrying infection due to their contact history or symptoms, to control the virus spread. Our study supports these conclusions and provides additional quantification of the amount of testing and tracing that would be needed to prevent a second wave of COVID-19 in the UK under different strategies to reopen schools and society from June 2020. To our knowledge, this is the first study to give such quantitative measures.

The model presented here has a number of limitations. Firstly, while we have made an effort to characterise the pandemic to resemble that of the UK, some of the parameters we have used are from a variety of sources across different settings as used in Covasim and outlined in Kerr et al.^12^ However, the main aspect we have focused on changing to illustrate different scenarios, is the transmission probability of social (household, school, work and community) contacts and the primary source for this was UK based.^15^ The changes we have simulated across scenarios reflect our understanding of possible options for school reopening as discussed in the UK. They are therefore fit for purpose within this analysis. Secondly, as with any modelling study, we have made a series of assumptions within the modelling framework. Specifically, we made assumptions about the proportion of COVID-19 infections that are symptomatic, as in the literature, there is a mixed evidence on this. While the World Health Organisation suggests that 80% of infections show mild symptoms^16^ and a recent study from the Italian city of Vo’ Euganeo at the epicentre of the European pandemic confirms that a large proportion, 50%-75%, of COVID-19 infections do not result in symptoms, other studies suggest this number is smaller: e.g. 10% among children,^17^ 18% among passengers on the Diamond Princess cruise ship^18^ and 42% among Japanese people returning from Wuhan, where the pandemic started.^19^ Changing this parameter in our model will change the transmission dynamics, and possibly change the peak of infection. The assumption in this study, as in Covasim, is that 70% of infection is symptomatic and guided by the findings by Davies et al.^22^ that the probability of developing clinical symptoms raises from around 20% in under 10s to over 70% in older adults. Future analyses will explore how changing the proportion of asymptomatic COVID-19 infections influences the impact of a TTI strategy, but this was beyond the remit of this study. Our assumption of a delay of one day to receive the test result and, once an individual tested positive, that they immediately isolated for 14 days may be slightly optimistic in the UK context. Finally, in the absence of robust data, our assumptions on infectiousness among children and young adults under 20 years old is based on an assumption and is varied in the sensitivity analysis. Future analysis of the virology of COVID-19 may suggest that infectiousness among children is even lower than 50%, although there are no data suggesting higher transmission than in adults.^5^ Our model can be rerun when further evidence becomes available.

Our model and analyses caution against early school and society reopening in the absence of a fully implemented TTI strategy. We show that school and society reopening in combination with TTI strategies is able to reduce R to below 1 and diminish it, and hence likely to prevent a secondary pandemic wave of COVID-19, control the transmission of SARS-CoV-2, and prevent a large number of COVID-19 deaths. This is true both of analyses assuming children transmit COVID-19 similarly to adults and those assuming a lower infectivity amongst children. In our modelling we have assumed that reopening schools is not a binary off-on switch, but instead that reopening schools would be accompanied by broader changes. School reopening would allow parents to go back to work, as part of reopening a proportion of businesses that are anticipated to be an important step in restarting the economic activity within the society. Specifically, we simulated increasing not only the school transmission, but also increased transmission within workplaces and the community that would arise as a result of reopening of school and society. The exact numbers representing these changes in this analysis are based on modelling assumptions, and the model can be rerun if more reliable numbers are available in future.

In summary, our findings suggest that reopening schools should be part of the next step of gradual relaxing of lockdown, but only if it is combined with a fully implemented TTI strategy with high coverage. It is currently unclear when the UK TTI strategy will achieve sufficient coverage. Such a strategy, to prevent onward transmission, could possibly comprise of virus testing for active infection in symptomatic individuals (i.e. PCR tests for SARS-CoV-2), followed by contact-tracing of individuals within the network of the infected person and isolation of individuals showing symptoms or diagnosed positive for infection. This would be an alternative to intermittent lockdown measures while we await an effective vaccine against SARS-CoV-2.

## Data Availability

Publicly available data were used for modelling in this study.

## Statement on data quality

Publicly available data was collated and used for modelling purposes.

## Contribution

JPG and RV came up with the idea of the study. JPG, CCK, RMS and DM developed the specific modelling framework, based on the Covasim model developed by CCK, RMS, DM and DK. CCK, RMS, DM, RK and JPG collated data for the parameters used. JPG ran the modelling analysis with input from CCK, RMS and DM. JPG, RV and CB defined the different scenarios in the UK context following conversations with Scientific Pandemic Influenza Modelling Group which gives expert advice to the UK Department of Health and Social Care and wider UK Government. JPG wrote the manuscript with input from CB, RV, CCK, RMS, DM and DK. All authors approved the final version.

## Acknowledgments

We would like to acknowledge Dr Edwin van Leeuwen (Public Health England), Dr Tim Colbourn (UCL), Dr William Waites (University of Edinburgh) and Dr Simone Sturniolo (Science and Technologies Facilities Council, UK) for their helpful conversations about different modelling approaches and Prof. Ruth Gilbert (UCL) for reading an earlier version of the paper.

## Footnotes

1 With TI, the daily testing probability *p_s_* is that of voluntary testing of symptomatic persons. With either of the TTI strategies, the daily testing probability *p_s_* is of active testing of symptomatic persons from June 2020.

2 The results do not change if we run a larger number of simulations and we tested 1,3,6,8, 10 and 20 simulations. The difference is that the noise in the simulations increases with increased size of simulations and this is why we chose six simulations for the figures here.

## References

1. World Health Organisation. Novel coronavirus - China. 12 Jan 2020. https://www.who.int/csr/don/12-january-2020-novel-coronavirus-china/en/ (accessed 12 May 2020.

2. Ferguson NM, Laydon D, Nedjati-Gilani G, et al. Report 9: Impact of non-pharmaceutical interventions (NPIs) to reduce COVID-19 mortality and healthcare demand. London: Imperial College, 2020.

3. COVID-19 educational disruption and response. 2020. https://en.unesco.org/themes/education-emergencies/coronavirus-school-closures (accessed 19 March 2020.

4. Viner RM, Russell SJ, Croker H, et al. School closure and management practices during coronavirus outbreaks including COVID-19: a rapid systematic review. Lancet Child Adolesc Health 2020.

5. Brurberg KD. The role of children in the transmission of SARS-CoV-2 (COVID-19), 1st update—a rapid review. Oslo: Norwegian Institute of Public Health; 2020.

6. (NCIRS) NCfIRaS. COVID-19 in schools - the experience in NSW. Sydney: NSW Government, 2020.

7. Bi Q, Wu Y, Mei S, et al. Epidemiology and transmission of COVID-19 in 391 cases and 1286 of their close contacts in Shenzhen, China: a retrospective cohort study. Lancet Infect Dis 2020.

8. Viner RM, Mytton O, Bonell C, et al. Susceptibility to COVID-19 amongst children and adolescents compared with adults: a systematic review and meta-analysis. *medRxiv preprint server* 2020.

9. Jones TC, Muhlemann B, Veith T, et al. An analysis of SARS-CoV-2 viral load by patient age. Berlin: Charité - Universitätsmedizin, 2020.

10. Coronavirus (COVID-19) in the UK: Update 6 May 2020. 6 May 2020 2020. https://coronavirus.data.gov.uk/?ga=2.41681382.554770926.1588844302-1722033634.1588620867 (accessed 6 May 2020.

11. Ministry of Education standards for suspension. 2020. https://cpd.moe.gov.tw/articleInfo.php?id=2568 (accessed 25 April 2020.

12. Kerr CC, et al. Covasim: an agent-based model for COVID-9 dynamics and suppression scenarios. *medRxiv preprint server* 2020.

13. Zhang J, Litvinova M, Liang Y, et al. Changes in contact patterns shape the dynamics of the COVID-19 outbreak in China. Science 2020.

14. Davies NG, Klepac P, Liu Y, et al. Age-dependent effects in the transmission and control of COVID-19 epidemics. *medRxiv preprint server* 2020.

15. Prem K, Cook AR, Jit M. Projecting social contact matrices in 152 countries using contact surveys and demographic data. PLoS Comput Biol 2017; 13(9): e1005697.

16. World Health Organisation. Coronavirus disease 2019 (COVID-19) Situation Report—46, 2020.

17. Qiu H, Wu J, Hong L, Luo Y, Song Q, Chen D. Clinical and epidemiological features of 36 children with coronavirus disease 2019 (COVID-19) in Zhejiang, China: an observational cohort study. Lancet Infect Dis 2020.

18. Mizumoto K, Kagaya K, Zarebski A, Chowell G. Estimating the asymptomatic proportion of coronavirus disease 2019 (COVID-19) cases on board the Diamond Princess cruise ship, Yokohama, Japan, 2020. Euro Surveill 2020; 25(10).

19. Nishiura H, Kobayashi T, Miyama T, et al. Estimation of the asymptomatic ratio of novel coronavirus infections (COVID-19). Int J Infect Dis 2020; 94: 154-5.

20. Shim E, Tariq A, Choi W, Lee Y, Chowell G. Transmission potential and severity of COVID-19 in South Korea. Int J Infect Dis 2020; 93: 339-44.

21. Cohen J, Kupferschmidt. Countries test tactics in ‘war’ against COVID-19. Science 2020; 367(6484):1287–1288

22. Davies NG, Klepac P, Liu Y, Prem K, Jit M, CMMID COVID-19 working group, Eggo R. Age-dependent effects in the transmission and control of COVID-19 epidemics. assessed 27/05/2020. https://www.medrxiv.org/content/10.1101/2020.03.24.20043018v1.full.pdf

